# Respiratory syncytial virus-associated hospitalisation in adults with comorbidities in two European countries

**DOI:** 10.1101/2023.08.31.23294884

**Authors:** Richard Osei-Yeboah, Caroline Johannesen, Amanda Marie Egeskov-Cavling, Junru Chen, Toni Lehtonen, Arantxa Urchueguía Fornes, John Paget, Thea K. Fischer, Xin Wang, Harish Nair, Harry Campbell PROMISE investigators

## Abstract

**Background:** Individuals with comorbidities are at increased risk of severe RSV infection. We estimated RSV-associated respiratory tract infection (RTI) hospitalisation among adults aged 45 years and older with comorbidities in Denmark and Scotland.

**Methods:** By analysing national hospital and virological data, we estimated annual average number and rate of RSV-associated hospitalisations by seven selected comorbidities and age during 2010-2018. We estimated rate ratios (RRs) of RSV-associated hospitalisation and 95% uncertainty ranges in comorbid adults versus the overall populations.

**Results:** In Danish adults aged 45y+, RSV-RTI hospitalisation rates ranged from 3.1 per 1000 individuals with asthma, to 19.4 per 1000 individuals per year with chronic kidney disease (CKD). In Scotland, the rate ranged from 2.4 per 1000 individuals per year with chronic liver disease (CLD), to 9.0 per 1000 individuals per year with chronic obstructive pulmonary disease (COPD). In both countries, we found over 2-4-fold increased risk of RSV hospitalisation in adults with COPD, ischemic heart disease (IHD), stroke and diabetes, and 1.5-3-fold increased risk for adults with asthma and 3-7-fold for those with CKD. RSV hospitalisation rates among adults aged 45-64y with COPD, asthma, IHD or CKD were higher compared with the overall population aged 65-74y.

**Conclusion:** Findings of this study provide important evidence for identifying risk groups and assisting health authorities in RSV vaccination policy making.

## Background

Respiratory syncytial virus (RSV) is a major cause of respiratory tract infections (RTI), and leads to about 245,000 and 158,000 hospitalisations annually in young children and adults aged 18 years and older respectively in EU countries, with a disproportionate burden occurring in infants and adults aged 65 years and older [1, 2]. Besides older age, co-morbidities are a key risk factor for RSV hospitalisation in adult population. Previous epidemiological studies show that comorbidities, e.g., chronic respiratory diseases, chronic heart diseases and diabetes, are prevalent among adult patients hospitalised with RSV-associated respiratory tract infection[3–6]. Moreover, chronic kidney diseases and cardiovascular diseases are found to be linked with risk of RSV severe illnesses and mortality[7–9]. A systematic review estimated a high incidence of RSV-associated ARI, and a high risk of mortality from hospitalised RSV among adults with comorbidities (e.g., cystic fibrosis, congestive heart failure, chronic obstructive pulmonary disease, or immunosuppression) [10]. However, estimates of RSV-associated ARI hospitalisation rates among comorbid adults are sparse in terms of geographical diversity [10, 11] and of the type of comorbidities being investigated [11], and none is available yet in the European region. One RSV vaccine product, i.e., Arexvy, has been authorised for use to prevent severe RSV-associated illnesses by European Medicines Agency [12]. In view of the rapid progress on development and evaluation of RSV vaccines and prophylaxis products, estimates of RSV hospitalisation rates in comorbid adults could provide timely evidence for recommendations on RSV immunisation among the adult population on the horizon.

Using routinely collected data from national hospital registries and virological surveillance, we aimed to estimate the number, rates and risk of RTI hospitalisations associated with RSV in pre COVID-19 era among Danish and Scottish adults aged 45 years and older that had at least one of seven comorbidities (specified below in case definitions), selected based on influenza vaccination recommendations and data availability, using a regression modelling approach [13]. Regression models are widely used to estimate RSV disease burden, especially among adults, while accounting for under-ascertainment of RSV diseases due to the lack of systematic RSV testing [14] and poor sensitivity of RSV-specific ICD-10 codes in routine clinical care practice [15, 16], and imperfect sensitivity of viral diagnostic tests [17], and the potentially poor detection of RSV in late-stage disease samples.

## Methods

### Study design and data sources

The study design and data sources have been described previously [18]. Briefly, we conducted a retrospective analysis using national hospital registries and virological surveillance data in Denmark (2010 to 2018) and Scotland (2010 to 2016). A season was from week 40 of one year to week 39 of the following year.

### Case definitions

As done previously [18, 19], we defined RTI hospitalisations based on ICD-10 diagnosis codes (Supplementary Table 1). RTI admission was defined as an admission with any mention of RTI in the diagnosis codes. We included seven comorbidities, i.e., chronic obstructive pulmonary disease (COPD), asthma, ischemic heart disease (IHD), stroke, diabetes, chronic kidney diseases (CKD) and chronic liver disease (CLD) according to Scotland recommendations on high-risk conditions for influenza vaccination and data availability[20]. The comorbidities were identified using ICD-10 diagnosis codes according to previous disease burden study (ICD-10 codes are given in Supplementary Table 2) [21]. We searched all the diagnostic fields to identify individuals that were hospitalised due to and with the diseases, and were recorded on any occasions during healthcare utilisation within 5 years before the RTI hospital episode or at the episode.

Scottish Burden of Disease group estimates of the prevalence of the seven chronic medical conditions in 2014 was used to derive hospitalisation rates in Scotland [21]. The prevalence of IHD, stroke, diabetes and CKD in Danish adults in 2015, and the prevalence of COPD and asthma in 2014, obtained from Danish disease burden reports and registers [22, 23] were used to derive hospitalisation rates in Denmark. Due to small case counts, Danish adults with CLD were excluded from the study.

### Statistical analyses

#### Model overview

Data were accessed and analysed separately by partners in Denmark and Scotland, based on the same analytical approach. A multiple linear regression model was used to estimate the number of RTI hospitalisations associated with RSV, as “RSV-associated RTI”, in adults aged 45y+ with chronic medical conditions similar to previous analyses [18, 24, 25]. Overall, the model included a natural cubic spline function for weeks during the study period, the number of RSV positive tests, and the number of influenza positive tests. In Scotland, we considered an interaction term of influenza-positive tests and season (2010-11 season; other seasons) as there were a greater number of influenza-positive tests in the 2010-11 season compared to other seasons, which may reflect changes in testing practices over the study period. We modelled separately for each of the chronic medical conditions in the two countries; for each condition, we further conducted subgroup analyses by age groups (45-54y, 55-64y, 65-74y, 75-84y, and 85y+) where appropriate. For each comorbidity and age group, we tested for the optimal lag/lead combination on RSV and influenza among a lag/lead of 0-3 weeks. The goodness of fit was assessed based on adjusted R-squared and the Akaike information criterion (AIC). Details of the model structure are given in Supplementary File 1.

We estimated the annual number of RSV-associated RTI hospital admissions based on model coefficients for RSV, and the number of RSV-positive tests. The 95% confidence intervals (CIs) were estimated using a 52-week-block bootstrap with 1 000 replicates. We estimated rate ratios (RRs) of RSV-associated RTI hospitalisation between the comorbid population and the overall population (with or without the comorbidity) of the same age band. The 95% uncertainty ranges (URs) of RR were estimated based on 1000 samples generated from a log-normal distribution of estimates of hospitalisation rates, with the 2.5^th^ percentile of the samples as the lower bound and the 97.5^th^ percentile of the samples as the upper bound as previously done [26].

### Sensitivity analyses

We conducted the following sensitivity analyses among adults aged 45y+ to assess the robustness of RSV burden estimates: (1) assuming zero lags for RSV and influenza predictors; (2) removing the influenza predictor from the models of which the coefficient was negative; (3) using the Poisson regression model; (4) time series data of rhinovirus positive tests were added to the models for Scottish population.

### Ethical statement

Data access approvals were obtained in both countries and analyses were conducted separately in each country in a secured environment.

## Results

### Average annual number and proportion of RSV–associated RTI hospitalisation in comorbid adults aged 45y+

We estimated that, in Danish adults aged 45y+ with comorbidities, the average annual number of RSV-associated RTI hospitalisations ranged from 165 (95% CI: 160 – 218) in adults with CKD to 1876 (1613 – 2178) in adults with COPD (Supplementary Table 3). RSV-associated RTI accounted for between 5.8% (4.5 – 7.3) in all RTI hospitalisations among adults with stroke and 16.8% (14.2 –19.4) among adults with asthma (Supplementary Table 4).

In Scottish adults aged 45y+ with comorbidities, the average annual number of RSV-associated RTI hospitalisations ranged from 64 (95% CI: 25 – 93) in adults with CLD to 914 (95% CI: 564 – 1122) in adults with IHD. The proportion of RSV-RTI hospitalisations ranged from 3.6% (1.4 – 5.0) in all RTI hospitalisations among adults with CLD to 7.4% (4.5 – 8.4) among adults with stroke (Supplementary Table 4). Estimates from the regression models generally fitted the observed data well (Supplementary Figure 1 and 2). In comparison, average annual estimates of influenza-associated RTI hospitalisation ranged from 77 (32– 80) among adults with CLD to 888 (371 – 1073) among adults with IHD (Supplementary Table 3).

### Average annual rate and rate ratio of RSV-associated RTI hospitalisation

RSV-RTI hospitalisation rates in the comorbid adults aged 45y+ varied between Denmark and Scotland (Table 1). In Denmark, we found the highest risk in adults with CKD, with a rate of 9.4 (95% CI: 18.9 - 25.7) RSV-RTI hospitalisations per 1 000 individuals per year, and an RR of 7.2 (95% UR: 5.2 - 10.3) compared with Danish overall population aged 45y+. For the other comorbidities, we estimated, approximately, a four-fold higher rate in Danish adults with COPD and IHD versus the overall population, a two-fold higher rate in those with diabetes and stroke, and a 1.5-fold higher rate in those with asthma. In Scotland, the highest risk was in adults with COPD, with a rate of 7.1 (4.6 – 8.8) RSV-RTI hospitalisations per 1 000 individuals per year, and an RR of 5.9 (95% UR 4.1 – 8.7) versus the overall population aged 45y+. For the remaining comorbidities, we estimated a four-fold higher rate in adults with IHD compared with the overall population, a three-fold higher rate in those with asthma, stroke and CKD, and a two-fold higher rate in those with CLD and diabetes (Table 1). In comparison, rates of influenza-associated RTI hospitalisation ranged from 4.4 (1.8 – 4.6) per 1000 adults with CLD per year to 7.6 (3.7 – 8.1) for those with asthma (Supplementary Table 3).

**Table 1.** Hospitalisation rates of RSV-RTI per 1 000 individuals per year in adults aged 45 years and older with selected underlying medical conditions, and rate ratios (RR) compared with the overall population.^1^.

|  | Denmark |  | Scotland |  |
| --- | --- | --- | --- | --- |
|  | Hospitalisation rate<br>(95% CI) | RR (95% UR) | Hospitalisation<br>rate (95% CI) | RR (95% UR) |
| **Overall population**<br>**(with or without**<br>**comorbidities)** | 2.0 (1.7,2.3) | Ref | 1.2 (0.8,1.4) | Ref |
| **COPD** | 9 (7.7,10.4) | 4.5 (3.7,5.5) | 7.1 (4.6,8.8) | 5.9 (4.1,8.7) |
| **Asthma** | 3.1 (2.6,3.6) | 1.5 (1.3,1.9) | 3.8 (2.6,4.8) | 3.2 (2.2,4.7) |
| **IHD** | 7.6 (6.4,8.8) | 3.8 (3.1,4.7) | 4.6 (2.8,5.6) | 3.8 (2.7,5.6) |
| **Stroke** | 3.7 (3.2,4.3) | 1.8 (1.5,2.3) | 3.5 (2.1,3.9) | 2.9 (2.0,4.3) |
| **Diabetes** | 4.7 (4.0,5.5) | 2.3 (1.9,2.9) | 2.4 (1.5,2.8) | 2.0 (1.4,2.9) |
| **Chronic kidney**<br>**disease** | 19.4 (18.9,25.7) | 9.7 (8.0,11.9) | 3.3 (2.1,3.7) | 2.7 (1.9,4.0) |
| **Chronic liver disease** | -- | -- | 2.4 (0.9,3.5) | 2.0 (1.4,2.9) |

\* COPD: chronic obstructive pulmonary disease. IHD: Ischemic heart disease. CKD: chronic kidney diseases, CLD: chronic liver disease, conditions were selected according to influenza vaccination recommendations and data availability.

An exploratory analysis shows that in Denmark, the estimated yearly RSV-RTI hospitalisation rates in comorbid adults aged 45y showed an increasing trend between the 2013/14 and 2017/18 seasons (Supplementary Figure 3). In the contrast, the rates remained similar in Scotland from the 2010/11 to 2015/16 seasons (Supplementary Figure 4).

### RSV-RTI hospitalisation rates in comorbid adults by age group

In the two nations, hospitalisation rates rose quite steeply from the 60s to 80s age groups, and were the highest in those aged 85y+ across most of comorbidity groups (Table 2). In particular, adults with COPD and asthma had more than six-fold higher hospitalisation rates at 75y+ compared with their younger counterparts in the two nations. The age-related patterns were less profound for the other comorbidities. Across age groups, the comorbid adults had higher rates compared to the overall population with an RR estimate above 1.0, except for several occasions (Figure 1 and 2, Supplementary Table 6). The RR estimates for comorbid adults versus the overall population were generally higher in those aged 45-54y and/or 55-64y (Figure 1 and 2). In particular, Danish adults and Scottish adults aged 45-54 years with IHD had an RR estimate of 7.8 (95% UR 6.0-10.3) and 6.0 (2.5,15.6); Scottish adults aged 55-64y with COPD had an RR estimate of 6.7 (3.7-12.7). Exceptions to this pattern were observed in adults with asthma (both countries) and stroke (Scotland only). In the two countries, adults with asthma had a particularly high RR estimate at 75y+; whereas Scottish adults with stroke had similar rates to the overall population at 45-64y. Danish adults with diabetes had an RR of 4.8 (3.7,6.3) at 45-54y, but a RR below 1.0 at 55y+; Scottish adults aged 65-74y and younger had an RR estimate above 1.0.

**Figure 1.**
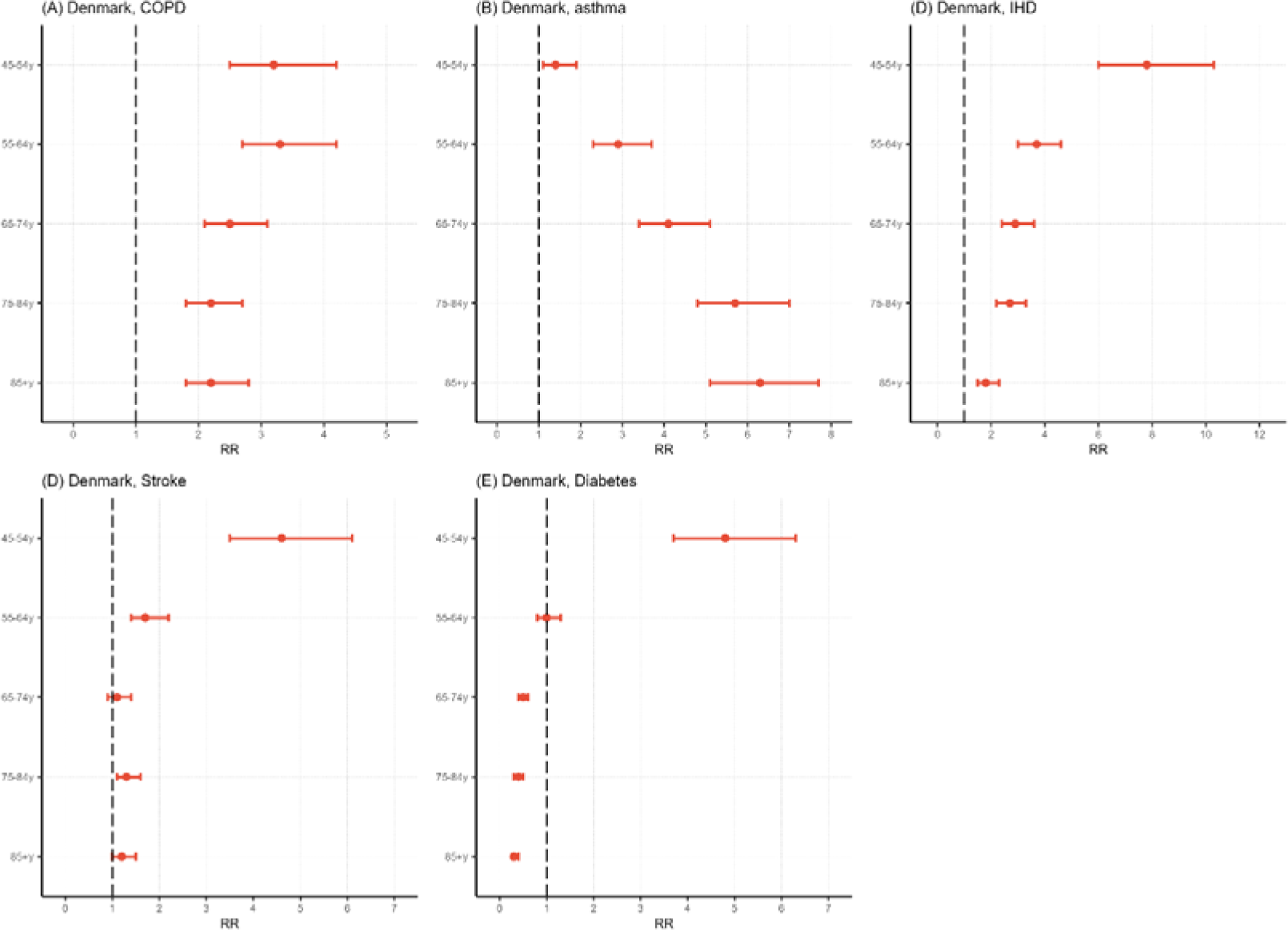
Age-specific rate ratios of RSV-associated RTI hospitalisation among the Danish adults versus Danish overall population. Panels show estimates for COPD (A), asthma (B), IHD (C), stroke (D), and diabetes (E).

**Figure 2.**
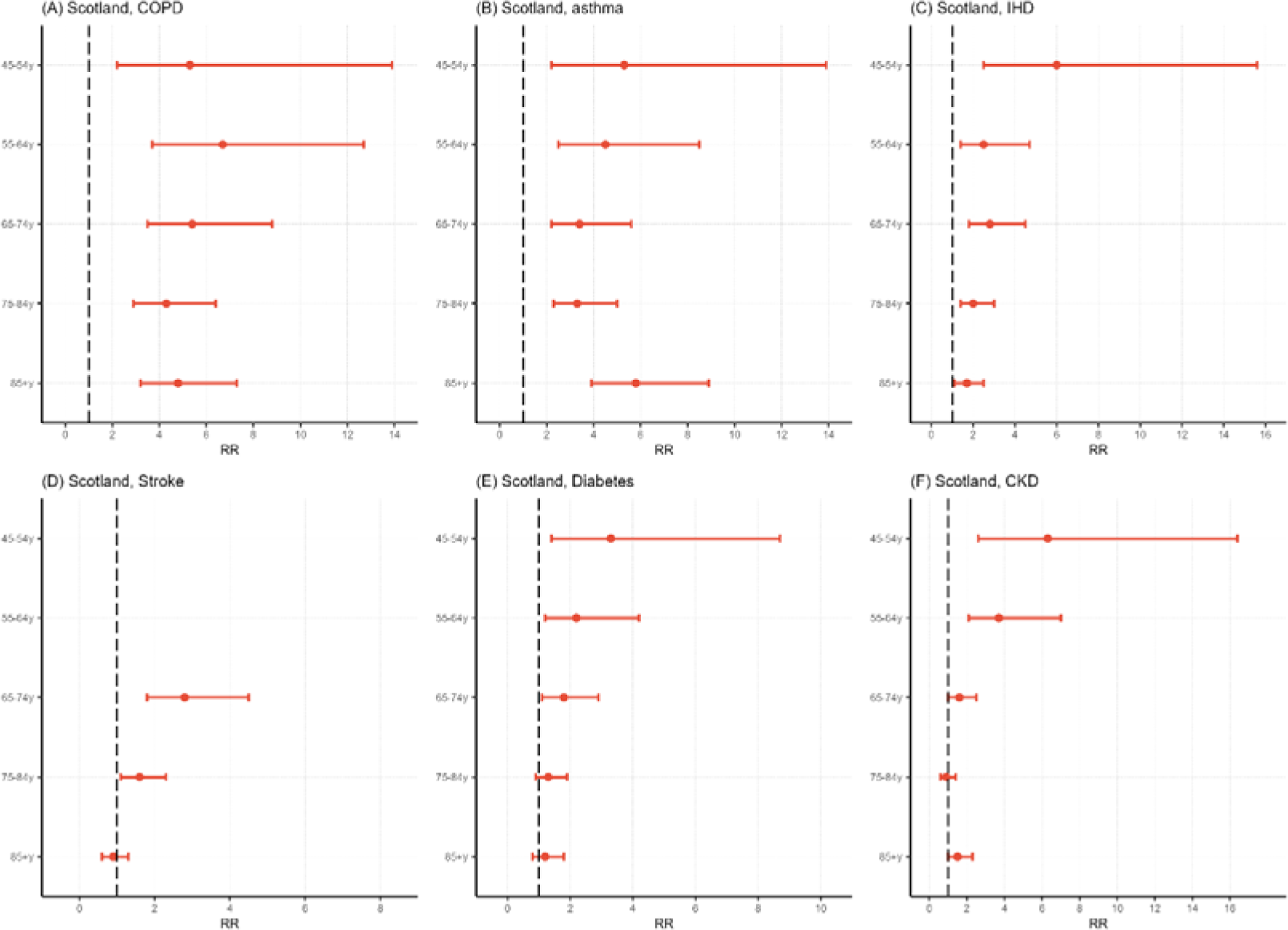
Age-specific rate ratios of RSV-associated RTI hospitalisation among the Scottish adults versus Scottish overall population. Panels show estimates for COPD (A), asthma (B), IHD (C), stroke (D), diabetes (E), and chronic kidney diseases (F). RR was not calculated for the 45-54y and 55-64y with stroke, as the lower bound of their rate estimates was negative.

**Table 2.** Hospitalisation rates of RSV-RTI per 1 000 individuals per year in adults aged 45 years and older with selected underlying medical conditions, by age groups.

|  | Overall population<br>(with or without<br>comorbidities) | COPD | Asthma | IHD | Stroke | Diabetes | Chronic kidney<br>disease<sup>†</sup> |
| --- | --- | --- | --- | --- | --- | --- | --- |
| **Denmark** |  |  |  |  |  |  |  |
| **45-54y** | 0.5 (0.4,0.6) | 1.6 (1.3,1.8) | 0.7 (0.6,0.8) | 3.9 (3.3,4.6) | 2.3 (1.9,2.6) | 2.4 (2.0,2.8) | -- |
| **55-64y** | 1.2 (1.0,1.4) | 4.0 (3.4,4.7) | 3.5 (3.0,4.1) | 4.4 (3.7,5.1) | 2.2 (1.8,2.5) | 1.2 (1.1,1.4) | -- |
| **65-74y** | 2.0 (1.7,2.3) | 5.1 (4.3,5.9) | 8.3 (7.0,9.7) | 5.9 (5.0,6.9) | 2.3 (1.9,2.6) | 1.0 (0.8,1.1) | -- |
| **75-84y** | 4.9 (4.2,5.6) | 10.5 (8.9,12.2) | 27.6 (23.4,31.9) | 12.9 (11.0,14.9) | 6.4 (5.5,7.5) | 1.9 (1.7,2.3) | -- |
| **85+y** | 7.9 (6.7,9.1) | 17.6 (14.9,20.4) | 49.5 (42.5,57.7) | 14.4 (12.0,16.7) | 9.4 (8.0,11.0) | 2.3 (2.2,3.1) | -- |
| **Scotland** |  |  |  |  |  |  |  |
| **45-54y** | 0.3 (0.1,0.4) | 1.6 (0.5,2.4) | 1.6 (0.8,2.2) | 1.8 (0.4,2.8) | 0.4 (-0.1,1.0) | 1.0 (0.4,1.6) | 1.9 (0.1,3.2) |
| **55-64y** | 0.4 (0.2,0.5) | 2.7 (1.8,3.5) | 1.8 (1.0,2.3) | 1.0 (0.4,1.6) | 0.8 (-0.3,1.7) | 0.9 (0.4,1.2) | 1.5 (0.9,2.1) |
| **65-74y** | 0.9 (0.6,1.2) | 4.9 (3.0,6.6) | 3.1 (1.4,5.1) | 2.5 (1.5,4.3) | 2.5 (1.3,3.7) | 1.6 (1.0,2.3) | 1.4 (0.9,1.7) |
| **75-84y** | 3.0 (2.0, 3.6) | 12.8 (8.3, 17.4) | 10.0 (2.9, 9.6) | 6.0 (3.8, 7.5) | 4.7 (2.4, 5.7) | 3.9 (2.0,4.7) | 2.8 (1.7, 3.3) |
| **85+y** | 8.2 (5.2,9.6) | 39.2 (21.9,47.5) | 47.9 (28,53.2) | 13.7 (8.1,17.5) | 7.2 (3.7,9.3) | 9.9 (5.8,12.5) | 12.3 (7.6,14.2) |

<sup>†</sup> We were unable to provide age-specific estimates for Scottish adults with CLD or for Danish adults with CKD, due to the small RTI case counts in the groups.

### Sensitivity analyses

Estimates of RSV-RTI hospitalisation rates in the sensitivity analyses were broadly comparable to the main analyses in two countries (Supplementary Table 7). Scottish estimates of rates in the comorbid adults aged 45y+, based on Poisson regression models and the models with addition of rhinovirus positive cases, changed by around 10% or less compared to those in the main models. Danish estimates for adults with COPD, IHD and asthma using Poisson regression models were about 13% to 17% lower than the main analyses; the estimates for the other comorbidities only changed marginally (<10% changes). Models with zero lags in RSV and influenza predictors showed lower accuracy compared with the main models, yielding comparable estimates for RSV in the Scottish population, and lower estimates in the Danish population.Discussion

Using national hospital and virological databases, this study provides estimates of RSV-associated RTI hospitalisation among adults aged 45 years and older with seven comorbidities, and their RRs versus the overall population (with or without comorbidities) in Scotland and Denmark. We found that in Danish adults aged 45y+, RSV-RTI hospitalisation rates ranged from 3.1 per 1000 individuals per year with asthma, to 19.4 per 1000 individuals per year with CKD; the rate ranged from 2.4 per 1000 individuals per year with CLD, to 9.0 per 1000 individuals per year with COPD in Scotland. Adults with the comorbidities showed a 1.5-fold to seven-fold higher rate compared to the overall populations aged 45y+ across two countries. Adults with COPD, IHD and diabetes consistently showed over four-, three- and two-fold higher rates in two countries, while more between-country differences were found for asthma and CKD. Across age groups, comorbid adults had higher rates compared to the overall populations of same age except for several occasions. Adults aged 65y+ with the comorbidities (except for Danish adults with diabetes) have a substantially high burden of RSV hospitalisation, especially among those with COPD or asthma.

Our estimates of RSV-RTI hospitalisation rates in adults with COPD are comparable to a US study that reported a rate of 13 per 1000 individuals per season in adults with COPD or chronic heart failure aged 65y+, while greater than the results by a New Zealand study [27]. In the New Zealand study, the RSV-RTI hospitalisation rate was between 0.2 and 1.4 per 1,000 individuals among adults aged 50y+ with COPD, asthma, congestive heart failure, coronary artery disease, cerebrovascular accident, diabetes, or end-stage renal disease. The disparities in estimates of RSV-RTI hospitalisation rates could be related to the use of different analytical approaches. In contrast to our study using a regression modelling approach, the New Zealand study reported laboratory-confirmed RSV hospital cases.

The RR estimates reflect the risk of RSV-RTI hospitalisation associated with the comorbidities, and are broadly comparable to previous population-based studies. A modelling study in England showed that adults aged 65y+ with a range of comorbidities were four-fold more likely be hospitalised for RSV-associated respiratory diseases compared to those without comorbidities [28]. In New Zealand, adults with seven comorbidities had between two-fold and 10-fold higher rates compared to those without the comorbidities, and the RR varied across age groups and comorbidities [11]. Similar to our results, the New Zealand study found a smaller increase in the risk of RSV-RTI hospitalisation in adults with diabetes compared to chronic respiratory and heart diseases, and the risk elevation was mainly observed in young and middle-aged adults with diabetes and was less notable in older adults[11]. Our RR estimates should be viewed as a conservative estimation of the risk in the comorbid adults in Denmark and Scotland, as we used the total population, with or without comorbidities, as a control.

The age-specific estimates of RSV hospitalisation rates and RR provide evidence on high-risk population groups that are relevant for RSV immunisation recommendations and participant selection for vaccine evaluation in clinical trials and real-world studies. Besides, using the overall adults aged 65-74y as a benchmark, estimates of RSV hospitalisation rates among adults with certain comorbidities remained elevated at 45-54y (COPD, asthma and IHD) or 55-64y (COPD, asthma, IHD and CKD). The point estimates in adults with CKD aged 45-54y were higher than the Scottish overall population aged 65y+, though it was difficult to draw a conclusion due to the wide confidence intervals of the estimates.

We found that RSV-associated RTI hospitalisation rates in Scottish adults aged 45y+ with the comorbidities were similar to influenza (Supplementary table 5). Findings of the comparison could assist health authorities in RSV vaccination policy making, in the presence of risk-based recommendations on influenza vaccination.

There are limitations and challenges in our study. One challenge in analyses of comorbid adults relates to the difficulty in defining and identifying comorbidities[11, 25]. In this study, we identified comorbidities that were recorded on any occasions in the national hospital-care registries within five years before an RSV hospital episode, by assuming that pre-existing comorbidities were recorded in any diagnostic fields at least once during recent healthcare utilisations prior to the RSV episode. It is possible that some individuals, especially adults at younger ages, were not diagnosed because they had been at early stage of disease progression, had mild symptoms or had remained good health status for five years or longer time. Given these factors and possible incomplete recording of comorbidities in the databases, our study provides a conservative estimate of true RSV-associated RTI hospital burden in adults with the comorbidities. Second, we assume that the virological data (RSV and influenza) in the overall population is an indicator of viral activity in the comorbid adults. The assumption is made based on observations that the time series data of hospitalised RTI cases and the virological data had similar trends and times of peaks (Supplementary Figure 1 and 2). Where exist, variations in RTI hospitalisation rates and potential temporal sequential patterns in viral circulation between the comorbidities and age groups have been accounted for, via the independent modelling process [25]. Moreover, the model fits and results from sensitivity analyses suggest that our estimates of RSV-RTI hospitalisation rates are generally robust. Third, we were unable to provide precise age-specific estimates for Scottish adults with CLD or Danish adults with CKD, due to the small RTI case counts in the groups. Finally, some of the individuals might have more than one comorbidity, which could cause biases in the estimates. Future research could compare the role of multiple comorbidities on risk of RSV infection. It is important to recognise the profound effect of COVID-19 and the impact of vaccination on RSV epidemiology and seasonality, which will likely confound any modern attempts to define RSV burden especially in high-risk populations.

## Conclusion

Using a standardised approach on national hospital and virological databases, we provide age-specific estimates of RSV-RTI hospitalisation among adults with seven comorbidities and aged 45y+ in two European countries. Our results show that adults aged 65y+ with these comorbidities, and adults aged 45-64y with COPD, IHD, asthma, or CKD remain a high priority population to consider for RSV immunisation when RSV vaccines become available for use in adult population [12, 29].

## Data Availability

All data produced in the present study are available upon reasonable request to the data controllers in both countries.

## Footnote Page

The PROMISE investigators are as follows:

Harry Campbell and Harish Nair (University of Edinburgh), Hanna Nohynek (THL, Finland), Anne Teirlinck (RIVM, The Netherlands), Michiel van Boven (RIVM, The Netherlands), Terho Heikkinen (University of Turku and Turku University Hospital, Finland), Louis Bont (University Medical Center Utrecht), Peter Openshaw (Imperial College, London), Phillipe Beutels (University of Antwerp), Andrew Pollard (University of Oxford), Veena Kumar (Novavax), Alexandro Orrico Sánchez (FISABIO, Spain), David Gideonse (RIVM, The Netherlands), Tin Tin Htar (Pfizer), Charlotte Vernhes (Sanofi Pasteur), Gael Dos Santos (GlaxoSmithKline), Rachel Cohen (GlaxoSmithKline), Jeroen Aerssens (Janssen), Rolf Kramer (Sanofi Pasteur), Ombeline Jollivet (Sanofi Pasteur), Nuria Manchin (TEAMIT).

## Author Contributions

HC and HN conceptualised the study. XW and ROY analysed the Scottish data. CJ and AEC analysed the Danish data. JC prepared figures and tables. ROY and XW led the interpretation and wrote the draft with important inputs from HN, HC, TL, AUF, JP, TKF, CJ, AEC and JC. All the authors critically reviewed the manuscript.

## Acknowledgements

We acknowledge the support of the electronic data research and innovation services (eDRIS) team at Public Health Scotland for their involvement in obtaining approvals, provisioning and linking, and the use of the secure analytical platform with the National Safe Haven.

