## Supplementary data for "Respiratory syncytial virus-associated hospitalisation in adults with comorbidities in two European countries"

**Supplementary file for “Respiratory syncytial virus-associated hospitalisation in adults with comorbidities in two European countries”**

**Model structure**

We used a multiple linear regression model to estimate the number of respiratory tract infection (RTI) hospital admissions associated with RSV in adults with chronic medical conditions similar to previous studies (1-3). Overall, the model included a natural cubic spline function for weeks during the study period, the number of RSV-positive tests, and the number of influenza-positive tests. In Scotland, we considered an interaction term of influenza-positive tests and season (2010-11 season; other seasons) as there were a greater number of influenza-positive tests in the 2010-11 season compared to other seasons, which may reflect changes in testing practices over the study period:

$$E(Y_{t,a})={\beta_{1,a}S}_{t}+\beta_{2,a}RSV_{t-ta1}+\beta_{3,a}Flu_{t-ta2}*2010/11season$$

t: week index (from 1 to the length of the study period)

a: Age group a

Y: weekly number of RTI hospital episodes (3-week moving average)

S: a natural cubic spline function of weeks.

RSV: weekly RSV positive tests

Flu: influenza-positive tests.

ta1: lags and/or leads in RSV

ta2: lags and/or leads in influenza.

In Denmark, the number of influenza A and influenza B positive tests was available and included in the model.

**Supplementary file 2**

Supplementary Table 1: The list of ICD-10 codes used to identify RTI diagnosis.

| **Diagnosis** | | **ICD-10** |
| --- | --- | --- |
| Acute upper respiratory tract infection (URTI) | | J00 J02.0 J02.8 J02.9 J03.0 J03.8 J03.9 J04.0 J04.1 J04.2 J05.0 J05.1 J06.0 J06.8 J06.9 |
| Lower respiratory tract infection (LRTI) | Pneumonia and influenza | J09 J10.0 J10.1 J10.8 J11.0 J11.1 J11.8 J12.0 J12.1 J12.2 J12.3 J12.8 J12.9 J13 J14 J15.0 J15.1 J15.2 J15.3 J15.4 J15.5 J15.6 J15.7 J15.8 J15.9 J16.0 J16.8 J17.0 J17.1 J17.2 J17.3 J17.8 J18.0 J18.1 J18.2 J18.8 J18.9 |
|  | Bronchiolitis and bronchitis | J20.0 J20.1 J20.2 J20.3 J20.4 J20.5 J20.6 J20.7 J20.8 J20.9 J21.0 J21.1 J21.8 J21.9 J40 |
|  | Unspecified LRTI | J22 |

Supplementary Table 2: Chronic comorbidities and ICD-10 codes (any diagnostic fields).

| **Condition** | **ICD-10 codes** |
| --- | --- |
| Chronic obstructive pulmonary disease | J40, J41.0, J41.1, J41.8, J42.0, J42.1, J42.4, J43.0, J43.1, J43.2, J43.8, J43.9, J44.0, J44.1, J44.8, J44.9, J47.0, J47.1, J47.9 |
| Asthma | J45.0, J45.1, J45.2, J45.3, J45.4, J45.5, J45.8, J45.9, J46 |
| Ischemic heart disease | I20.0, I20.1, I20.8, I20.9, I21.0, I21.1, I21.2, I21.3, I21.4, I21.6, I21.9, I22.0, I22.1, I22.2, I22.8, I22.9, I23.0, I23.1, I23.2, I23.3, I23.4, I23.5, I23.6, I23.7, I23.8, I24.0, I24.1, I24.8, I24.9, I25.0, I25.1, I25.2, I25.3, I25.4, I25.5, I25.6, I25.7, I25.8, I25.9 |
| Stroke | I63.0, I63.1, I63.2, I63.3, I63.4, I63.5, I63.6, I63.7, I63.8, I63.9, I65.0, I65.1, I65.2, I65.3, I65.8, I65.9, I66.0, I66.1, I66.2, I66.3, I66.4, I66.8, I66.9, I67.2, I67.3, I67.5, I67.6, I69.3, G45.0, G45.1, G45.2, G45.3, G45.4, G45.8, G45.9, G46.0, G46.1, G46.2, G46.3, G46.4, G46.5, G46.6, G46.7, G46.8. |
| Diabetes | E10.0, E10.1, E10.3, E10.4, E10.5, E10.6, E10.7, E10.8, E10.9, E11.0, E11.1, E11.3, E11.4, E11.5, E11.6, E11.7, E11.8, E11.9, E12.0, E12.1, E12.3, E12.4, E12.5, E12.6, E12.7, E12.8, E12.9, E13.0, E13.1, E13.3, E13.4, E13.5, E13.6, E13.7, E13.8, E13.9, E14.0, E14.1, E14.3, E14.4, E14.5, E14.6, E14.7, E14.8, E14.9, P70.0, P70.1, P70.2, R73.0, R73.9 |
| Chronic kidney disease | D63.1, E10.2 , E11.2 , E12.2, E13.2 , E14.2, I12.0, I12.1, I12.2, I12.9, I13.0, I13.1, I13.2, I13.9, N02.0, N02.1, N02.2, N02.3, N02.4, N02.5, N02.6, N02.7, N02.8, N02.9, N03.0, N03.1, N03.2, N03.3, N03.4, N03.5, N03.6, N03.7, N03.8, N03.9, N04.0, N04.1, N04.2, N04.3, N04.4, N04.5, N04.6, N04.7, N04.8, N04.9, N05.0, N05.1, N05.2, N05.3, N05.4, N05.5,N05.6, N05.7, N05.8, N05.9, N06.0, N06.1, N06.2, N06.3, N06.4, N06.5, N06.6, N06.7, N06.8, N06.9, N07.0, N07.1, N07.2, N07.3, N07.4, N07.5, N07.6, N07.7, N07.8, N07.9, N08.0, N08.1, N08.2, N08.3, N08.4, N08.5, N08.8, N15.0, N18.0, N18.1, N18.2, N18.3, N18.4, N18.5, N18.6, N18.8, N18.9, Q61.0 Q61.1, Q61.2, Q61.3, Q61.4, Q61.5, Q61.8, Q61.9, Q62.0, Q62.1, Q62.2, Q62.3 ,Q62.4, Q62.5, Q62.6, Q62.7, Q62.8 |
| Chronic liver disease | I85.0, I85.9, K70.0, K70.1, K70.2, K70.3, K70.4, K70.9, K71.0, K71.1, K71.3, K71.4, K71.5, K71.7, K71.8, K71.9, K72.0, K72.1, K72.9, K73.0, K73.1, K73.2, K73.8, K73.9, K74.0, K74.1, K74.2, K74.3, K74.4, K74.5, K74.6, K74.9, K75.8, K75.9, K76.0, K76.6, K76.7, K76.9 |

Supplementary Table 3: Average annual number of RTI hospital admissions associated with viruses in the overall population and with selected comorbidities, by age group.

|  | The average annual number of RTI hospital admissions attributable to RSV and influenza virus (95% CI) | | | | | | | |
| --- | --- | --- | --- | --- | --- | --- | --- | --- |
|  | Overall | COPD | Asthma | IHD | Stroke | Diabetes | CKD | CLD |
| Denmark | | | | | | | | |
| **45+y** |  |  |  |  |  |  |  |  |
| RSV | 4975.9  (4225.8, 5785.9) | 1876.2  (1612.6,2178.4) | 433.4  (368, 502) | 1229.4  (1039, 1433) | 328.2  (282, 378) | 1032  (876, 1201) | 165  (160, 218) | -- |
| **45-54y** |  |  |  |  |  |  |  |  |
| RSV | 378.7  (322.2, 442.3) | 65.4  (55.8, 77) | 39.9  (33.5, 46.9) | 76.8  (64.4, 89.4) | 21  (18.0, 24.2) | 78.5  (65.6, 91.0) | -- | -- |
| **55-64y** |  |  |  |  |  |  |  |  |
| RSV | 845.0  (712.6, 980.3) | 265  (223.4, 308.2) | 143.7  (122.8, 167.4) | 177.1  (148, 204) | 38.7  (32.8, 44.8) | 71.7  (61.3, 83.3) | -- | -- |
| **65-74y** |  |  |  |  |  |  |  |  |
| RSV | 1214.9 (1029.2,1402) | 422.5  (360.1, 490.4) | 233.3  (196, 271.4) | 301.7  (255, 352) | 57.3  (48.4, 66.4) | 67.5  (57.3, 78.3) | -- | -- |
| **75-84y** |  |  |  |  |  |  |  |  |
| RSV | 1499.2  (1284.7, 1724.4) | 660.2  (560.7, 768) | 294.3  (248.9, 340) | 479  (408, 555) | 146.9  (125.4, 171.3) | 83.4  (71.1, 97.1) | -- | -- |
| **85+y** |  |  |  |  |  |  |  |  |
| RSV | 919.1  (780.2, 1056.9) | 258.9  (219.5, 299.8) | 153.5  (131.8, 178.9) | 203.2  (170, 235) | 113.9  (96.0, 132.2) | 38.7  (32.7, 45) | -- | -- |
| Scotland | | | | | | | | |
| **45+y** |  |  |  |  |  |  |  |  |
| Flu | 2293.2  (137.4, 3332.2) | 793.6  (392.7, 897.6) | 481.1  (235.5, 512.1) | 888.3  (371.2, 1073.0) | 258.3  (115.7, 326.2) | 579.2  (252.2, 721.9) | 489.7  (195.2, 643.0) | 77.1  (32.2, 80.4) |
| RSV | 28851.6  (1873.6, 3360.7) | 808.4  (526.1, 995.1) | 458.8  (319.3, 582.0) | 914.1  (563.9, 1122.4) | 314.8  (192.6, 357.4) | 588.0  (385.1, 701.1) | 559.2  (358.8, 621.3) | 63.9  (25.2, 93.2) |
| **45-54y** |  |  |  |  |  |  |  |  |
| Flu | 392.6  (28.7, 382.8) | 56.2  (23.6, 59.7) | 101.1  (63.6, 112.6) | 35.4  (3.3, 36.2) | 5.7  (-.8, 9.9) | 53.4  (30.7, 59.5) | 16.4  (5.3, 24.1) | -- |
| RSV | 200.9  (104.5, 299.2) | 26.0  (7.4, 39.3) | 70.6  (33.1, 95.0) | 32.3  (7.9, 49.0) | 3.4  (-.8, 7.7) | 40.0  (17.9, 66.9) | 15.4  (0.7, 25.9) | -- |
| **55-64y** |  |  |  |  |  |  |  |  |
| Flu | 499.7  (23.0, 472.4) | 130.6  (61.9, 131.5) | 97.1  (55.9, 95.3) | 113.3  (79.1, 116.9) | 30.6  (9.1, 35.2) | 87.8  (23.8, 111.3) | 40.3  (18.7, 50.2) | -- |
| RSV | 236.9  (137.4, 336.4) | 85.6  (57.0, 111.2) | 65.5  (35.2, 83.2) | 40.3  (14.2, 63.6) | 11.7  (-4.3, 26.4) | 56.1  (24.3, 74.1) | 35.3  (22.1, 50.9) | -- |
| **65-74y** |  |  |  |  |  |  |  |  |
| Flu | 637.7  (296.9, 648.1) | 217.3  (106.9, 238.6) | 115.8  (44.8, 131.1) | 227.5  (131.7, 241.5) | 53.0  (20.3, 77.9) | 166.9  (82.3, 182.1) | 92.3  (37.6,104. 9) | -- |
| RSV | 178.9  (165.7, 189.5) | 174.6  (107.9, 237.7) | 79.3  (35.1, 132.6) | 145.0  (88.0, 253.0) | 59.5  (32.1, 89.9) | 119.0  (70.8, 167.7) | 77.2  (48.3, 88.8) | -- |
| **75-84y** |  |  |  |  |  |  |  |  |
| Flu | 800.3  (342.9, 1037.3) | 247.9  (90.9, 346.9) | 96.9  (37.2, 118.1) | 314.4  (100.2, 449.4) | 85.3  (43.6, 126.6) | 174.2  (18.4, 274.2) | 170.8  (40.5, 244.4) | -- |
| RSV | 933.7  (621.3, 1122.2) | 314.3  (202.5, 425.6) | 128.8  (74.3, 187.1) | 354.9  (224.3, 447.3) | 129.7  (66.4, 157.6) | 214.6  (112.9, 258.9) | 178.3  (106.3, 211.4) | -- |
| **85+y** |  |  |  |  |  |  |  |  |
| Flu | 647.8  (228.9, 986.3) | 148.3  (53.8, 238.9) | 56.7  (11.8, 81.1) | 251.8  (87.0, 386.6) | 94.5  (39.1, 153.3) | 97.2  (10.0, 156.3) | 18.0  (70.9,262.4) | -- |
| RSV | 915.1  (577.7, 1068.8) | 203.2  (113.3, 245.9) | 105.7  (61.8, 117.4) | 337.4  (199.8,433.1) | 110.9  (56.4, 143.8) | 151.3  (88.6, 190.4) | 245.5  (151.3,283. 7) | -- |

Supplementary Table 4: Proportion of RTI hospital admissions that are associated with RSV and influenza virus (for Scotland), in the overall population and with selected underlying medical conditions, by age group and condition.

|  | The proportion of RTI admission attributable to RSV and influenza virus, % (95% CI) | | | | | | | |
| --- | --- | --- | --- | --- | --- | --- | --- | --- |
|  | Overall | COPD | Asthma | IHD | Stroke | Diabetes | CKD | CLD |
| **Denmark** | | | | | | | | |
| **45+y** |  |  |  |  |  |  |  |  |
| RSV | 10.8 (9.2,12.6) | 12.6 (10.8,14.6) | 16.8 (14.2,19.4) | 13.1 (11.1,15.3) | 5.8 (4.5,7.3) | 13.5 (11.5,15.7) | 11.6 (11.2,15.3) | -- |
| **45-54y** |  |  |  |  |  |  |  |  |
| RSV | 10.7 (9.1,12.5) | 10.7 (9.1,12.6) | 9.3 (7.8,11.0) | 27.4 (23.0,31.9) | 15.7 (13.5,18.1) | 23.2 (19.4,26.9) | -- | -- |
| **55-64y** |  |  |  |  |  |  |  |  |
| RSV | 13.0 (10.9,15.0) | 13.8 (11.6,16.0) | 13.8 (11.8,16.0) | 19.3 (16.1,22.2) | 8.4 (7.1,9.7) | 15.0 (12.8,17.4) | -- | -- |
| **65-74y** |  |  |  |  |  |  |  |  |
| RSV | 10.3 (8.8,11.9) | 9.8 (8.4,11.4) | 10.2 (8.6,11.9) | 12.8 (10.8,14.9) | 4.6 (3.9,5.4) | 9.5 (8.1,11.0) | -- | -- |
| **75-84y** |  |  |  |  |  |  |  |  |
| RSV | 10.7 (9.2,12.3) | 12.1 (10.3,14.1) | 11.4 (9.7,13.2) | 13.8 (11.7,16.0) | 7.9 (6.7,9.2) | 11.7 (9.9,13.6) | -- | -- |
| **85+y** |  |  |  |  |  |  |  |  |
| RSV | 9.0 (7.7,10.4) | 9.9 (8.4,11.4) | 11.7 (10.1,13.7) | 5.7 (7.3,10.1) | 8.0 (6.8,9.3) | 11.2 (9.5,13.1) | -- | -- |
| **Scotland** | | | | | | | | |
| **45+y** |  |  |  |  |  |  |  |  |
| Influenza | 6.6 (3.1,7.5) | 6.0 (2.9,6.7) | 7.6 (3.7,8.1) | 5.7 (2.4,6.9) | 6.1 (2.7,7.7) | 6.7 (2.9,8.4) | 6.1 (2.4,8.1) | 4.4 (1.8,4.6) |
| RSV | 6.4 (4.2,7.5) | 6.1 (3.9,7.5) | 7.3 (5.1,9.2) | 5.9 (3.6,7.3) | 7.4 (4.5, 8.4) | 6.8 (4.5,8.1) | 7.0 (4.5,7.8) | 3.6 (1.4,5.0) |
| **45-54y** |  |  |  |  |  |  |  |  |
| Influenza | 9.0 (0.7,8.8) | 7.8 (3.3,8.3) | 10.5 (6.6,11.7) | 6.4 (0.6,6.5) | 4.6 (-.7,8.0) | 9.9 (5.7,11.0) | 7.9 (2.6,11.6) | -- |
| RSV | 4.6 (2.4,6.9) | 3.6 (1.0,5.5) | 7.3 (3.4,9.9) | 5.8 (1.4,8.9) | 2.8 (-.7,6.3) | 7.4 (3.3,12.3) | 7.4 (0.3,12.5) | -- |
| **55-64y** |  |  |  |  |  |  |  |  |
| Influenza | 7.8 (0.4,7.4) | 7.4 (3.5,7.5) | 8.8 (5.0,8.6) | 7.5 (5.2,7.7) | 9.3 (2.8,10.7) | 7.7 (2.1,9.7) | 7.6 (3.5,9.5) | -- |
| RSV | 3.7 (2.2,5.3) | 4.9 (3.2,6.3) | 5.9 (3.2,7.5) | 2.7 (0.9,4.2) | 3.6 (-1.3,8.0) | 4.9 (2.1,6.5) | 6.7 (4.2,9.6) | -- |
| **65-74y** |  |  |  |  |  |  |  |  |
| Influenza | 6.4 (3.0,6.5) | 5.9 (2.9,6.5) | 8.2 (3.2,9.3) | 6.5 (3.8,6.9) | 6.1 (2.3,9.0) | 7.5 (3.7,8.2) | 6.8 (2.8,7.8) | -- |
| RSV | 4.7 (3.2,6.1) | 4.8 (2.9,6.5) | 5.6 (2.5,9.4) | 6.1 (5.7,6.5) | 6.9 (3.7,10.4) | 5.4 (3.2,7.5) | 5.7 (3.6,6.6) | -- |
| **75-84y** |  |  |  |  |  |  |  |  |
| Influenza | 5.9 (2.5,7.6) | 5.2 (1.9,7.2) | 5.4 (2.1,6.6) | 5.5 (1.7,7.8) | 5.3 (2.7,7.8) | 5.5 (0.6,8.6) | 5.6 (1.3,8.0) | -- |
| RSV | 6.8 (4.6, 8.2) | 6.6 (4.2, 8.9) | 7.2 (4.1, 10.4) | 6.2 (3.9, 7.8) | 8.0 (4.1, 9.7) | 6.8 (3.6,8.2) | 5.9 (3.5,7.0) | -- |
| **85+y** |  |  |  |  |  |  |  |  |
| Influenza | 6.3 (2.2,9.7) | 6.2 (2.2,10.0) | 5.5 (1.2,7.9) | 6.1 (2.1,9.3) | 7.3 (3.0,11.8) | 6.3 (0.6,10.1) | 6.4 (2.5.9.2) | -- |
| RSV | 9.0 (5.7,10.5) | 8.5 (4.7,10.3) | 10.3 (6.0,11.5) | 8.1 (4.8,10.4) | 8.6 (4.3,11.1) | 9.8 (5.7,12.3) | 8.6 (5.3,9.9) | -- |

Supplementary Table 5: Average annual hospital admission rates of influenza-RTI per 1000 adults aged 45 years and older with selected underlying medical conditions, and rate ratios (RR) compared with the overall population in Scotland.

|  | Overall population | COPD | | Asthma | | IHD | | Stroke | | Diabetes | | Chronic kidney disease | | Chronic liver disease | |
| --- | --- | --- | --- | --- | --- | --- | --- | --- | --- | --- | --- | --- | --- | --- | --- |
|  | Hospital admission rate (95% CI) | Hospital admission rate (95% CI) | RR (95% UR) | Hospital admission rate (95% CI) | RR (95% UR) | Hospital admission rate (95% CI) | RR (95% UR) | Hospital admission rate (95% CI) | RR (95% UR) | Hospital admission rate (95% CI) | RR (95% UR) | Hospital admission rate (95% CI) | RR (95% UR) | Hospital admission rate (95% CI) | RR (95% UR) |
| **Scotland** | | | | | | | | | | | | | | | |
| **45+y** |  |  |  |  |  |  |  |  |  |  |  |  |  |  |  |
| Flu-RTI | 1.2  (0.6,1.4) | 7.0  (3.5,7.9) | 5.8  (3.4,10.5) | 4.0  (1.9,4.2) | 3.3  (1.9,6.0) | 4.4  (1.9,5.4) | 3.7  (2.1,6.6) | 2.9  (1.3,3.6) | 2.4  (1.4,4.3) | 2.3  (1.0,2.9) | 1.9  (1.1,3.4) | 2.9  (1.2,3.8) | 2.4  (1.4,4.3) | 2.9  (1.2,3.0) | 2.4  (1.4,4.3) |
| **45-54y** |  |  |  |  |  |  |  |  |  |  |  |  |  |  |  |
| Flu-RTI | 0.5  (0.0,0.5) | 3.4  (1.4,3.6) | 6.7  (0.3,162) | 2.3  (1.5,2.6) | 4.5  (0.2,109.6) | 2.0  (0.2,2.1) | 3.9  (0.2,95.3) | 0.7  (-0.1,1.2) | NA | 1.3  (0.7,1.4) | 2.5  (0.1,61.9) | 2.0  (0.7,3.0) | 3.9  (0.2,95.3) | 2.2  (0.0,3.2) | NA |
| **55-64y** |  |  |  |  |  |  |  |  |  |  |  |  |  |  |  |
| Flu-RTI | 0.7  (0.0,0.7) | 4.1  (2.0,4.2) | 5.7  (0.2,175.9) | 2.6  (1.5,2.6) | 3.6  (0.2,111.6) | 2.9  (2.0,3.0) | 4.1  (0.2,124.4) | 2.0  (0.6,2.3) | 2.8  (0.1,85.8) | 1.4  (0.4,1.8) | 2.0  (0.1,60.1) | 1.7  (0.8,2.1) | 2.4  (0.1,72.9) | 2.3  (-0.8, 2.9) | NA |
| **65-74y** |  |  |  |  |  |  |  |  |  |  |  |  |  |  |  |
| Flu-RTI | 1.2  (0.6,1.2) | 6.1  (3.0,6.7) | 5.1  (3.3,8.2) | 4.5  (1.7,5.1) | 3.7  (2.4,6.0) | 3.9  (2.2,4.1) | 3.2  (2.1,5.2) | 2.2  (0.8,3.2) | 1.8  (1.2,3.0) | 2.3  (1.1,2.5) | 1.9  (1.2,3.1) | 1.7  (0.7,2.0) | 1.4  (0.9,2.3) | 4.3  (2.3, 5.7) | 3.6  (2.3,5.8) |
| **75-84y** |  |  |  |  |  |  |  |  |  |  |  |  |  |  |  |
| Flu-RTI | 2.6  (1.1,3.3) | 10.1  (3.7,14.2) | 3.9  (1.9,8.3) | 7.6  (2.9,9.2) | 2.9  (1.4,6.2) | 5.3  (1.7,7.6) | 2.0  (1.0,4.3) | 3.1  (1.6,4.6) | 1.2  (0.6,2.5) | 3.2  (0.3,5.0) | 1.2  (0.6,2.6) | 2.7  (0.6,3.8) | 1.0  (0.5,2.2) | 3.2  (-0.4, 4.6) | NA |
| **85+y** |  |  |  |  |  |  |  |  |  |  |  |  |  |  |  |
| Flu-RTI | 5.8  (2.1,8.8) | 28.6 (10.4,46.1) | 4.9  (2.0,13.2) | 25.7  (5.3,36.8) | 4.4  (1.8,11.9) | 10.2  (3.5,15.7) | 1.7  (0.7,4.7) | 6.1  (2.5,9.9) | 1.0  (0.4,2.8) | 6.4  (0.7,10.3) | 1.1  (0.4,3.0) | 9.1  (3.6,13.1) | 1.6  (0.6,4.2) | 10.2  (-9.5, 14.4) | NA |

Supplementary Table 6: Age-specific rate ratios (RR) and 95% uncertain ranges (UR) of RSV-RTI hospitalisation compared with the overall population.

|  | COPD | Asthma | IHD | Stroke | Diabetes | CKD |
| --- | --- | --- | --- | --- | --- | --- |
| Denmark |  |  |  |  |  |  |
| 45-54y | 3.2  (2.5,4.2) | 1.4  (1.1,1.9) | 7.8  (6.0,10.3) | 4.6  (3.5,6.1) | 4.8  (3.7,6.3) | -- |
| 55-64y | 3.3  (2.7,4.2) | 2.9  (2.3,3.7) | 3.7  (3.0,4.6) | 1.7  (1.4,2.2) | 1.0  (0.8,1.3) | -- |
| 65-74y | 2.5  (2.1,3.1) | 4.1  (3.4,5.1) | 2.9  (2.4,3.6) | 1.1  (0.9,1.4) | 0.5  (0.4,0.6) | -- |
| 75-84y | 2.2  (1.8,2.7) | 5.7  (4.8,7.0) | 2.7  (2.2,3.3) | 1.3  (1.1-1.6) | 0.4  (0.3,0.5) | -- |
| 85y+ | 2.2  (1.8,2.8) | 6.3  (5.1,7.7) | 1.8  (1.5,2.3) | 1.2  (1.0,1.5) | 0.3  (0.3,0.4) | -- |
| **Scotland** |  |  |  |  |  |  |
| 45-54y | 5.3  (2.2,13.9) | 5.3  (2.2,13.9) | 6.0  (2.5,15.6) | NA | 3.3  (1.4,8.7) | 6.3  (2.6,16.4) |
| 55-64y | 6.7  (3.7,12.7) | 4.5  (2.5,8.5) | 2.5  (1.4,4.7) | NA | 2.2  (1.2,4.2) | 3.7  (2.1,7.0) |
| 65-74y | 5.4  (3.5,8.8) | 3.4  (2.2,5.6) | 2.8  (1.8,4.5) | 2.8  (1.8,4.5) | 1.8  (1.1,2.9) | 1.6  (1.0,2.5) |
| 75-84y | 4.3  (2.9,6.4) | 3.3  (2.3,5.0) | 2.0  (1.4,3.0) | 1.6  (1.1,2.3) | 1.3  (0.9,1.9) | 0.9  (0.6,1.4) |
| 85y+ | 4.8  (3.2,7.3) | 5.8  (3.9,8.9) | 1.7  (1.1,2.5) | 0.9  (0.6,1.3) | 1.2  (0.8,1.8) | 1.5  (1.0,2.3) |

Supplementary Table 7: Sensitivity analyses for annual hospital admission rate of RSV-RTI per 1000 adults aged ≥45 years with selected comorbidities.

|  | | | ≥45 years | | | | | | |
| --- | --- | --- | --- | --- | --- | --- | --- | --- | --- |
|  |  |  | Comorbidity | | | | | | |
| Denmark | Sensitivity description | Virus | COPD | Asthma | IHD | Stroke | Diabetes | CKD | CLD |
|  | Main model | RSV | 9.0 (7.7,10.4) | 3.1 (2.6,3.6) | 7.6 (6.4,8.8) | 3.7 (3.2,4.3) | 4.7 (4.0,5.5) | 19.4 (18.9,25.7) | -- |
|  |  | AIC | 4312.8 | 3068.1 | 3948.7 | 3413.1 | 3734.0 | 2717.7 | -- |
|  | Assuming no lags for RSV and Flu | RSV | 6.5 (5.5,7.6) | 2.1 (1.8,2.4) | 5.0 (4.3,5.8) | 3.9 (3.3,4.4) | 2.9 (2.4,3.3) | 14.8 (12.5,17.1) | -- |
|  |  | AIC | 4373.5 | 3781.1 | 3984.1 | 3425.8 | 3083.8 | 2738.7 | -- |
|  | Use Poisson regression model | RSV | 7.5 (6.3,8.8) | 2.7 (2.3,3.2) | 6.5 (5.5,7.5) | 3.7 (3.1,4.3) | 4.4 (3.7,5.2) | 21.1 (17.6,25.0) | -- |
|  |  | AIC | 5781.8 | 3167.2 | 4469.3 | 3485.8 | 4071.7 | 2739.4 | -- |
| Scotland | Main model | Influenza | 7.1 (4.6,8.8) | 3.8 (2.6,4.8) | 4.6 (2.8,5.6) | 3.5 (2.1,3.9) | 2.4 (1.5,2.8) | 3.3(2.1,3.7) | 2.4 (0.9,3.5) |
|  |  | RSV | 7.0 (3.5,7.9) | 4.0 (1.9,4.2) | 4.4 (1.9,5.4) | 2.9 (1.3,3.6) | 2.3 (1.0,2.9) | 2.9 (1.2,3.8) | 2.9 (1.2,3.0) |
|  |  | AIC | 2692.9 | 2365.0 | 2776.7 | 2094.1 | 2513.0 | 2462.0 | 1679.6 |
|  | Assuming no lags for RSV and Flu | Influenza | 5.0 (4.6,5.2) | 2.7 (2.5,2.9) | 3.3 (3.1,3.5) | 2.6 (2.4,2.7) | 1.5 (1.4,1.6) | 2.3 (2.2,2.5) | 1.8 (1.7,1.9) |
|  |  | RSV | 6.4 (3.7,9.0) | 3.5 (2.0,4.9) | 4.0 (2.3,5.6) | 2.6 (1.5,3.7) | 2.1 (1.2,2.9) | 2.6 (1.5,3.6) | 2.9 (1.7,4.0) |
|  |  | AIC | 2915.9 | 2551.1 | 3011.1 | 2286.8 | 2764.6 | 2703.8 | 1724.33 |
|  | Add rhinovirus to model | Influenza | 6.0 (5.5,6.4) | 3.2 (2.9,3.4) | 3.8 (3.5,4.1) | 3.2 (2.9,3.4) | 2.0 (1.9,2.2) | 2.8 (2.6,3.0) | 1.9 (1.7,2.0) |
|  |  | RSV | 7.2 (3.6,10.9) | 4.0 (1.9,6.2) | 4.6 (2.2,7.0) | 2.9 (1.4,4.5) | 2.4 (1.1,3.7) | 3.0 (1.3,4.6) | 2.9 (1.7,4.1) |
|  |  | AIC | 2660.9 | 2330.0 | 2742.4 | 2071.4 | 2487.8 | 2432.0 | 1669.8 |
|  | Use Poisson regression model | Influenza | 7.0 (6.2,7.7) | 3.7 (3.2,4.3) | 4.4 (3.9,4.9) | 3.3 (2.9,3.8) | 2.3 (1.9,2.7) | 3.2 (2.6,3.7) | 2.4 (2.1,8.0) |
|  |  | RSV | 6.7 (3.2,10.4) | 3.8 (1.7,6.0) | 4.3 (1.9,6.7) | 2.7 (1.3,4.3) | 2.2 (0.9,3.5) | 2.7 (1.1,4.5) | 2.7 (1.5,4.0) |
|  |  | AIC | 2702.8 | 2370.8 | 2788.3 | 2120.8 | 2504.3 | 2452.6 | 1790.4 |

*COPD-Chronic obstructive pulmonary disease; IHD-Ischemic heart disease; CKD-Chronic kidney disease; CLD-Chronic liver disease

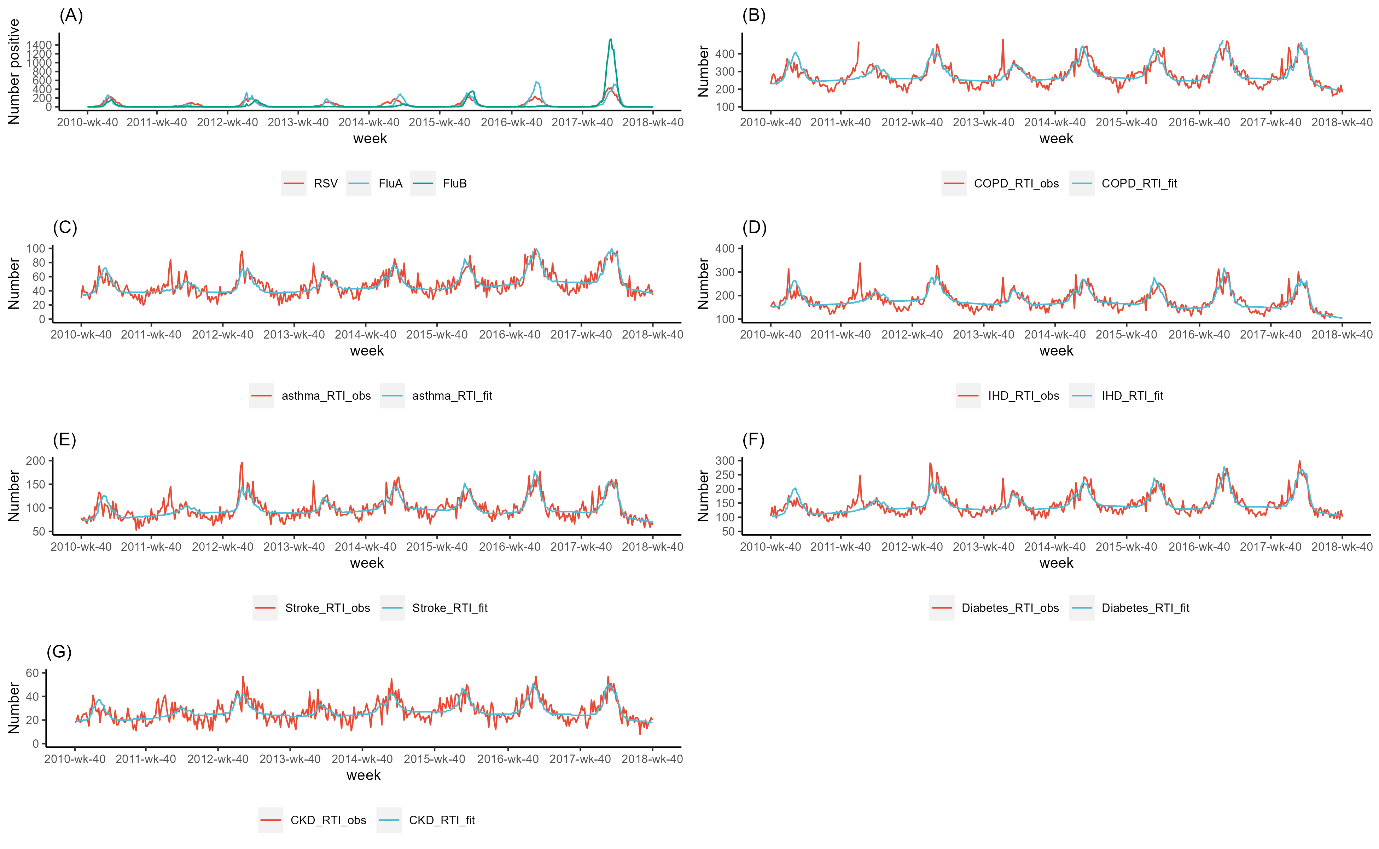

**Supplementary Figure 1.** Weekly time series for respiratory syncytial virus (RSV) and influenza virus (Panel A), and weekly time series of observed and fitted respiratory tract infection (RTI) hospitalisations in Danish adults aged 45 years and older with comorbidities, i.e., COPD (B), asthma (C), ischemic heart disease (D), stroke (E) diabetes (F), and chronic kidney disease (G).

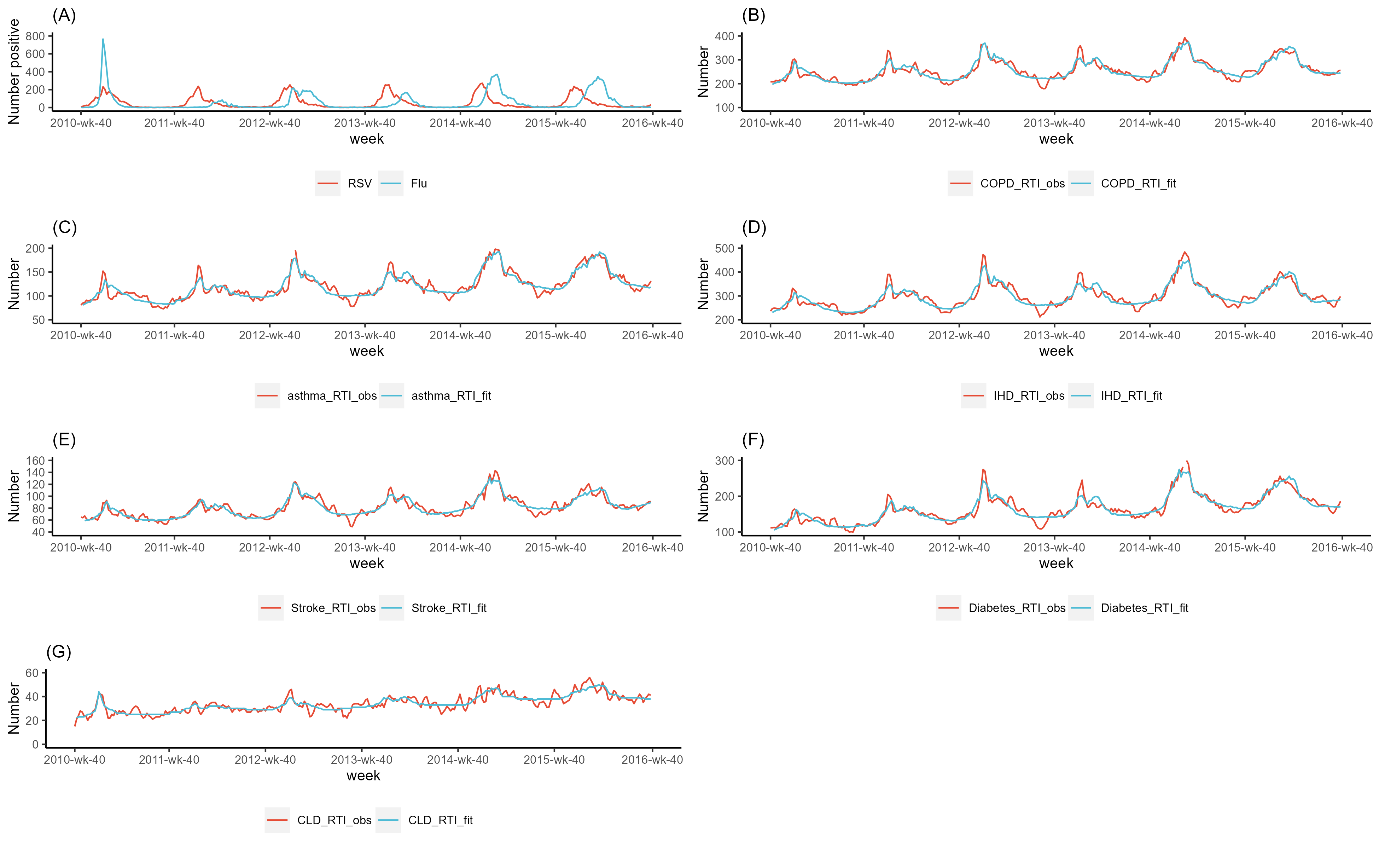

**Supplementary Figure 2.** Weekly time series for respiratory syncytial virus (RSV) and influenza virus (panel A), and weekly time series of observed (3-week moving average) and fitted respiratory tract infection (RTI) hospitalisations in Scottish adults aged 45 years and older with comorbidities, i.e., COPD (B), asthma (C), ischemic heart disease (D), stroke (E), diabetes (F), and chronic liver disease (G).

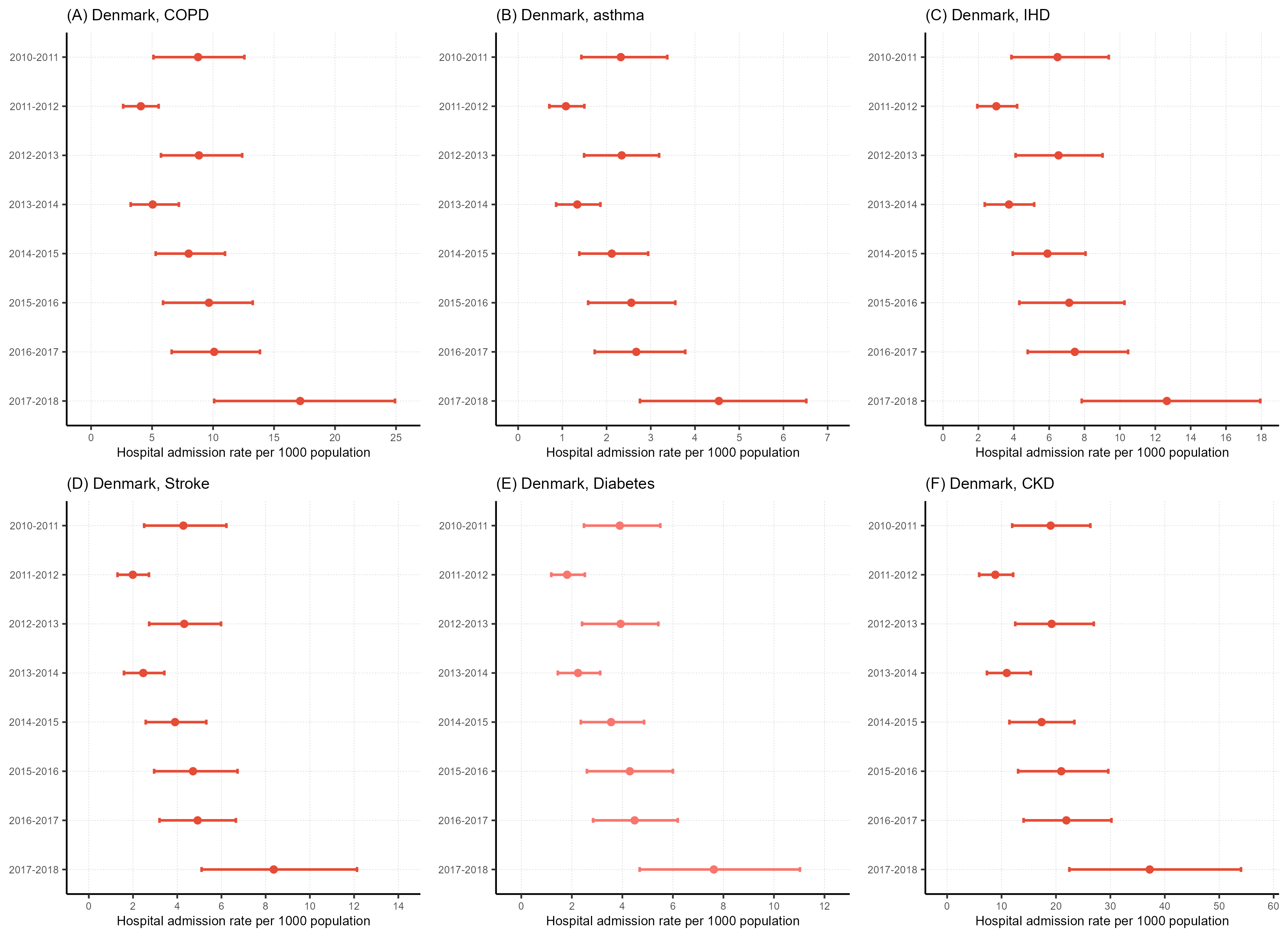

**Supplementary Figure 3**. Annual hospitalisation rates of RSV-RTI per 1000 persons in Danish adults aged 45 years and older with underlying medical conditions, by condition and season (2010-2018), i.e., COPD (A), asthma (B), IHD (C), stroke (D) diabetes (E), and chronic kidney disease (F).

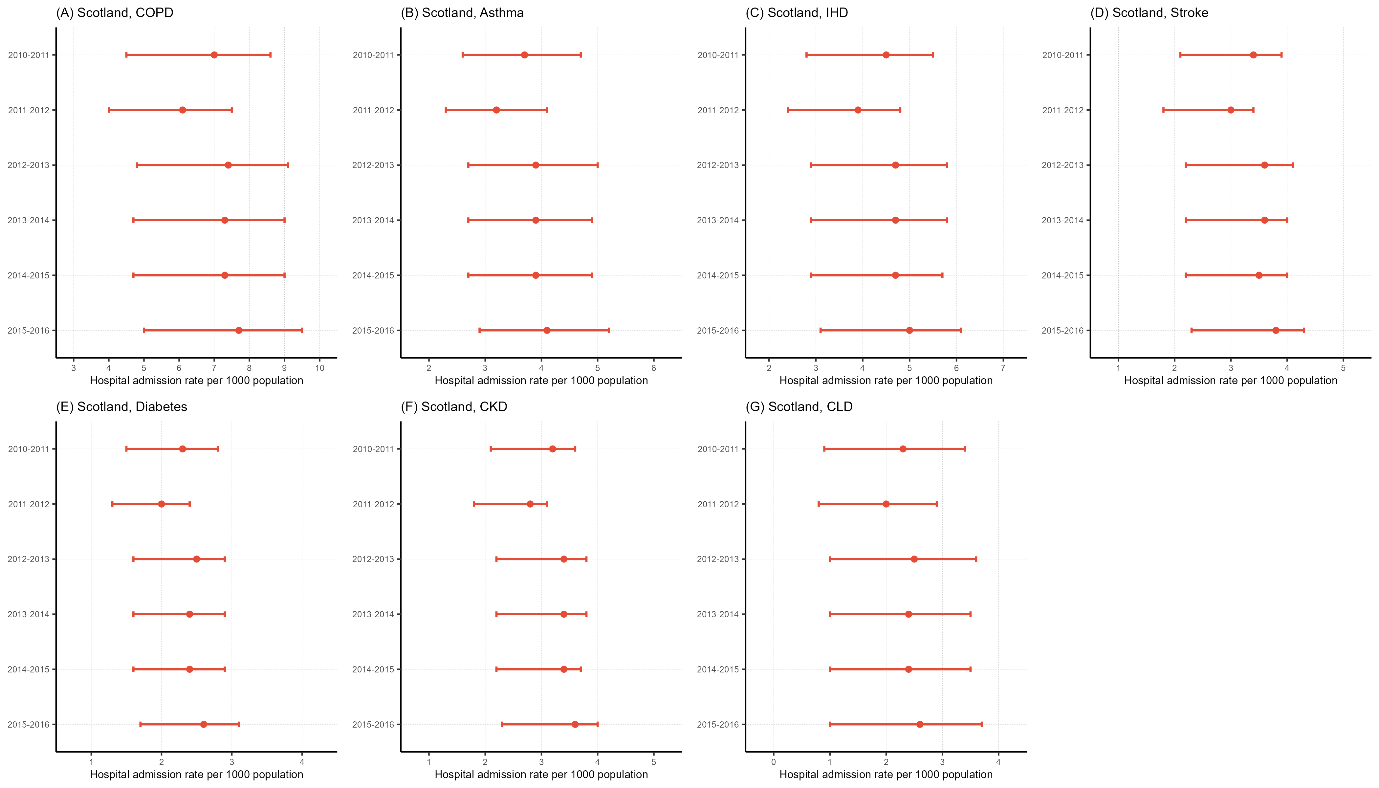

**Supplementary Figure 4.** Annual hospitalisation rates of RSV-RTI per 1000 persons in Scottish adults aged 45 years and older with underlying medical conditions, by condition and season (2010-2016), i.e., COPD (A), asthma (B), IHD (C), stroke (D), diabetes (E), chronic kidney disease (F) and chronic liver disease (G).
